# Implementation of Point of Care Ultrasound in extended focused assessment with sonography for trauma at a rural hospital in Uganda

**DOI:** 10.1101/2023.12.21.23300400

**Authors:** Rita Nassanga, Sam Bugeza, Ameda Faith, Irene Kabanda, Harriet Nakiberu, Tadeo Semaganda, Francis Olweny, Roy Nagawa, Aloysius Gonzaga Mubuuke

## Abstract

**Introduction:** Focused Assessment with Sonography for Trauma (FAST) is a rapid bedside ultrasound examination performed at presentation of a trauma patient whereas the extended FAST (eFAST) examines each hemithorax for the presence of free fluid and air. It is an ‘extension’ of the trauma clinical assessment process and aids rapid diagnosis with an aim of identifying free peritoneal fluid which allows for immediate transfer to theatre or further imaging. FAST can be performed by surgeons, emergency physicians, and paramedics as a screening test to detect post-traumatic pericardial effusion or hemoperitoneum, performed at presentation of a trauma patient.

**Objective:** The purpose of this study was to evaluate the feasibility of implementing a point of care ultrasound training in trauma for non-imaging health professionals and evaluate the ability of the trainees to apply the knowledge and skills gained to perform the ultrasound examination among trauma patients.

**Methods:** It was a prospective cohort study conducted at Kiwoko hospital, a rural based hospital in Uganda. The study involved developing a curriculum and training of clinicians in point of care ultrasound for trauma patients through didactic lectures and practical sessions as well as assessing them at baseline and thereafter further assessment at three months follow up.

**Results:** A total of 19 clinicians were initially enrolled however, 12 were evaluated at baseline and nine were followed up for three months. The median length of time in clinical practice of the clinicians in this study was 11(2-36) months. At baseline, majority of the clinicians correctly identified and named all the anatomical structures pertinent to images obtained in eFAST; sub-xiphoid view 7(58.3%), right upper quadrant view 5(41.7%), left upper quadrant view 6(50.0%), suprapubic view 7(58.3%), thoracic-pleural fluid view 7(58.3%), and thoracic-pneumothorax view 4(33.3%). At follow-up, the clinicians demonstrated acceptable competency in ultrasound technique when performing eFAST in most views assessed. However, compared to the baseline observation, a statistically significant decline (p=0.001) in image quality was noted in evaluation of organs in the left upper quadrant.

**Conclusion:** Clinicians generally demonstrated acceptable competency in performing eFAST assessment in trauma patients. With more training, frequent hands-on practice, regulation and adequate supervision, clinicians can ably perform eFAST procedures to aid in management of trauma patients.

## INTRODUCTION

Focused Assessment with Sonography for Trauma (FAST) is a rapid point-of-care ultrasound (POCUS) examination performed at the time of presentation of a trauma patient[1]. The use of POCUS was first described in 1988 in emergency medicine[2]. The extended FAST (eFAST) examines each hemithorax for the presence of free fluid and air[1]. FAST aids rapid diagnosis through identifying free peritoneal fluid, assumed to be haemopaeritoneum in the context of trauma which if found, allows for immediate transfer to theatre for life saving interventions [3, 4]. FAST has an overall excellent specificity [3, 5]and high sensitivity for occult pneumothoraces[6]. It is invariably performed by a formally trained clinician and is considered an ‘extension’ of the trauma clinical assessment process, to detect post-traumatic pericardial effusion or hemoperitoneum, and pneumothorax performed at the time of presentation of a trauma patient for quick management.

Trauma is among the leading causes of death and disability in the world[7]. In Africa, road traffic death rates account for 26.6/100,000 people. In Uganda, where this study was conducted, road traffic injuries, mainly due to motorcyclists (41 % of accidents) are the leading cause of abdominal trauma and the third leading cause of hospital admissions and mortality[3]. There are 24 people killed per 100 road crashes and on average, Uganda loses 10 people per day in road traffic accidents making it the highest level in East Africa[7]. Before FAST/eFAST, invasive procedures such as peritoneal lavage and exploratory laparotomy were commonly utilized to diagnose visceral injury[4]. FAST/eFAST in trauma aids immediate diagnosis, is non-invasive, readily available and plays a pivotal role in the initial diagnosis of trauma-related visceral injuries, but remains under-utilized especially in low resource contexts[6]. POCUS shows significant improvement in performance of ultrasound by physicians, following hands on practical training[8]. Despite known diagnostic values, the utilisation of POCUS in emergencies is low and is proportional to the level of clinical expertise among the users thus training and utility of POCUS among clinicians and trainees should be further advocated and supported[9].The number of imaging specialists in many low resource settings is low and many of them are concentrated in urban tertiary hospitals. Thus, despite the presence of ultrasound equipment in many rural based health facilities, there are no trained personnel to operate them leading to delayed or missed diagnosis.

In Uganda, the Ministry of Health started supplying ultrasound equipment to district level hospitals and Health Centers IVs, but this equipment remains under-utilized due to lack of trained imaging personnel, yet it would be used to conduct rapid assessment of trauma patients. Therefore, the purpose of the study was to evaluate the feasibility of implementing a point of care ultrasound training in trauma for non-imaging health professionals and evaluate the ability of the trainees to apply and retain the knowledge and skills gained to perform the ultrasound examination among trauma patients.

## Materials and Methods

### Design and setting

This was a prospective cohort study conducted from 20^th^ September 2022 to 30^th^ May 2023, at Kiwoko hospital, a rural based hospital in Uganda. The study involved training of clinicians in eFAST and then following them up to assess their competency in applying the knowledge and skills gained after three months. Kiwoko hospital has a bed capacity of 250, acts as a referral health facility for the community catchment area and receives a number of trauma related cases daily. It has several cadres of health workers including surgeons, medical officers, imaging technologists, nurses and midwives. There are only two imaging technologists who conduct all the imaging related examinations.

### Study population

The participants involved clinical officers, medical officers and surgeons who were purposively selected. These were initially trained in conducting eFAST because they directly manage trauma patients. All those who had ever received POCUS training were excluded from the study. A total of nineteen participants were selected for this training.

### The eFAST training curriculum

A team of experts in radiology designed the curriculum with well-defined targeted outcomes to be acquired. The outcomes were related to identification free fluid collection which is a common among trauma patients. The four basic competency areas where trainees were expected to demonstrate free fluid collection in trauma patients were: perihepatic space (including Morison’s pouch or the hepatorenal recess), perisplenic space, pericardium, and the pelvis. The trainees underwent two didactic lectures for four weeks to acquire knowledge about ultrasound and the examinations they were expected to perform. These lectures focused on basic knowledge of Ultrasound Physics, Knobology of the ultrasound scan machine, standard technique of systematically performing an ultrasound scan, key trans-abdominal scanning planes of the abdomen, probe orientation and proper probe selection. They were also trained on how to identify major abdominal organs, free fluid collection in the abdomen and pleural cavities, free air in the chest cavity as well as professionalism and ethical conduct including appropriate time to refer the patient for further radiology assessment.

For practical sessions, one healthy participant was selected among the hospital employees and the other was an ambulant patient with mild ascites and mild pleural effusion. Demonstrations were carried out by the training experts as the trainees observed. The experts demonstrated the standard technique and thereafter allowed the trainees to perform the ultrasound examination with close supervision.

Demonstrations focused on both normal findings and pathology so that trainees got acquainted with both.

### Data management and Statistical analysis

Data in this study was analyzed using STATA statistical software version 14. Descriptions were made using frequencies and percentages. Comparison was made using Fisher’s exact test.

### Ethical Considerations

Ethical approval to conduct the study was granted by the School of Medicine Research Ethics Committee (Mak-SOMREC-2022-307). All study participants provided written informed consent to participate in the study. Confidentiality of all participants was ensured and data was only accessible to the researchers.

## Results

Nineteen clinicians were initially trained and all attended the first didactic lecture on eFAST skills transfer at Kiwoko Hospital. Only 16 attended the second didactic eFAST practical hands on training whereas 12 underwent baseline eFAST skills evaluation. However, only nine clinicians were finally evaluated on their eFAST skills three months later following the training. Table 1 illustrates the different cadres initially trained.

**Table 1:**
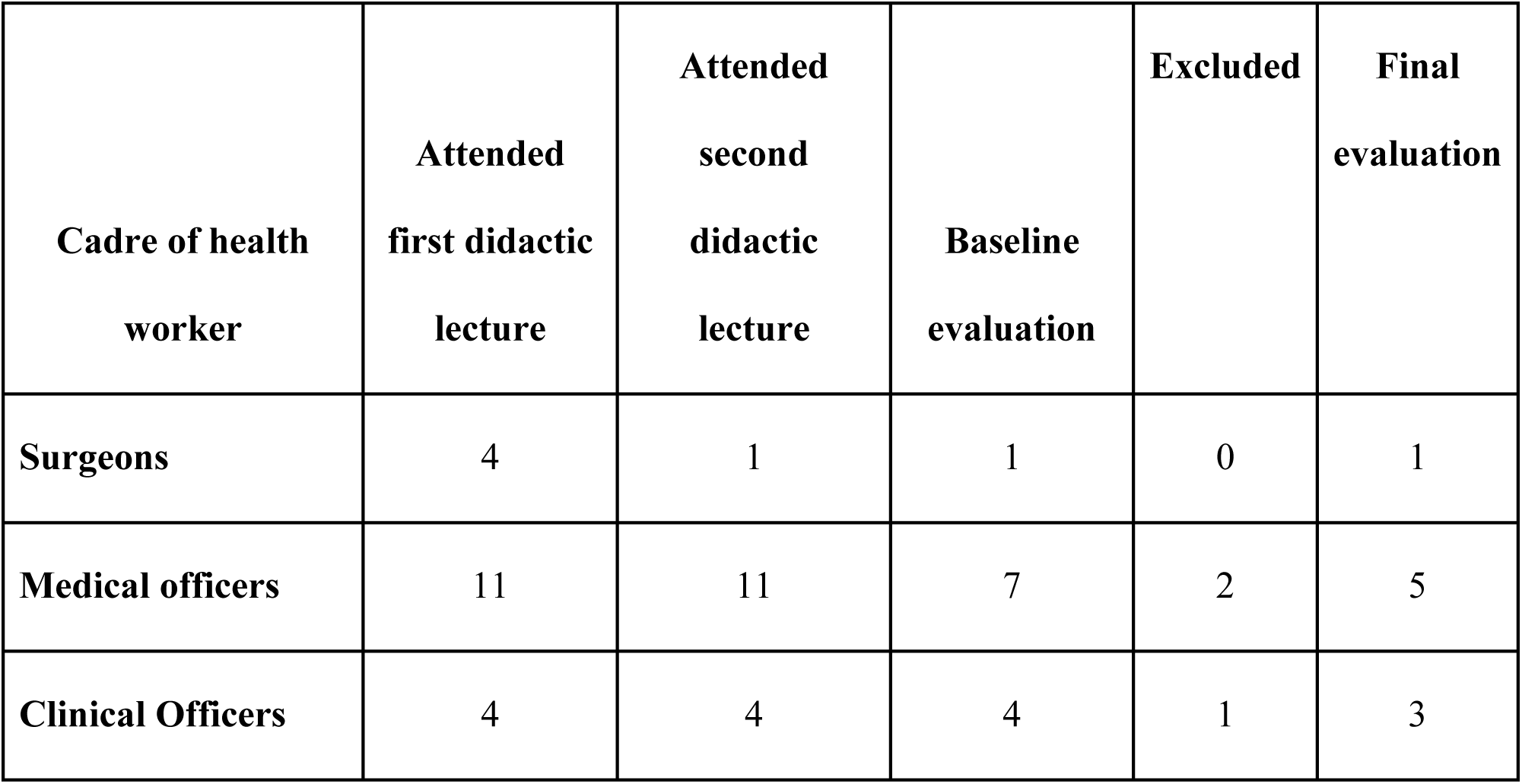
Cadres of health workers at Kiwoko Hospital during study timeline.

| **Cadre of health worker** | **Attended first didactic lecture** | **Attended second didactic lecture** | **Baseline evaluation** | **Excluded** | **Final evaluation** |
| --- | --- | --- | --- | --- | --- |
| **Surgeons** | 4 | 1 | 1 | 0 | 1 |
| **Medical officers** | 11 | 11 | 7 | 2 | 5 |
| **Clinical Officers** | 4 | 4 | 4 | 1 | 3 |

### Baseline assessment of the trainees

The baseline assessment was carried out two weeks post-training Clinicians demonstrated good skills with the technique used in ultrasound scan in performing eFAST in each of the eFAST views assessed. About 91.7%(n=11) of the 12 clinicians correctly positioned the probe and oriented selection marker on the sub-xiphoid eFAST view. Furthermore, majority of the clinicians obtained either outstanding images with no suggestions for improvements or excellent images with minor suggestions for improvement in each of the five eFAST views. For instance, in sub-xiphoid view, 5 (41.7%) clinicians obtained outstanding quality images with no suggestions for improvement and an equal number of clinicians-5 (41.7%) also obtained quality excellent images with minor suggestions for improvement.

Generally, majority of the clinicians correctly identified and named all the anatomical structures pertinent to images obtained in eFAST; sub-xiphoid view 7(58.3%), right upper quadrant (RUQ) view 5(41.7%), left upper(LUQ) quadrant view 6(50.0%), suprapubic view 7(58.3%), thoracic-pleural fluid view 7(58.3%), and thoracic-pneumothorax view 4(33.3%). Clinicians also majorly correctly acquired interpreted images to answer all relevant eFAST questions in; sub-xipoid view 6(50.0%), right upper quadrant view 5(41.7%), left upper quadrant view 5(41.7%), suprapubic view 7(58.3%), thoracic-pleural fluid view 6(50.0%),. The clinicians were also able to interpret some but not all relevant eFAST questions in thoracic-pneumothorax view 4(33.3%). Table 2 summarizes the baseline evaluation of the trainees.

**Table 2:**
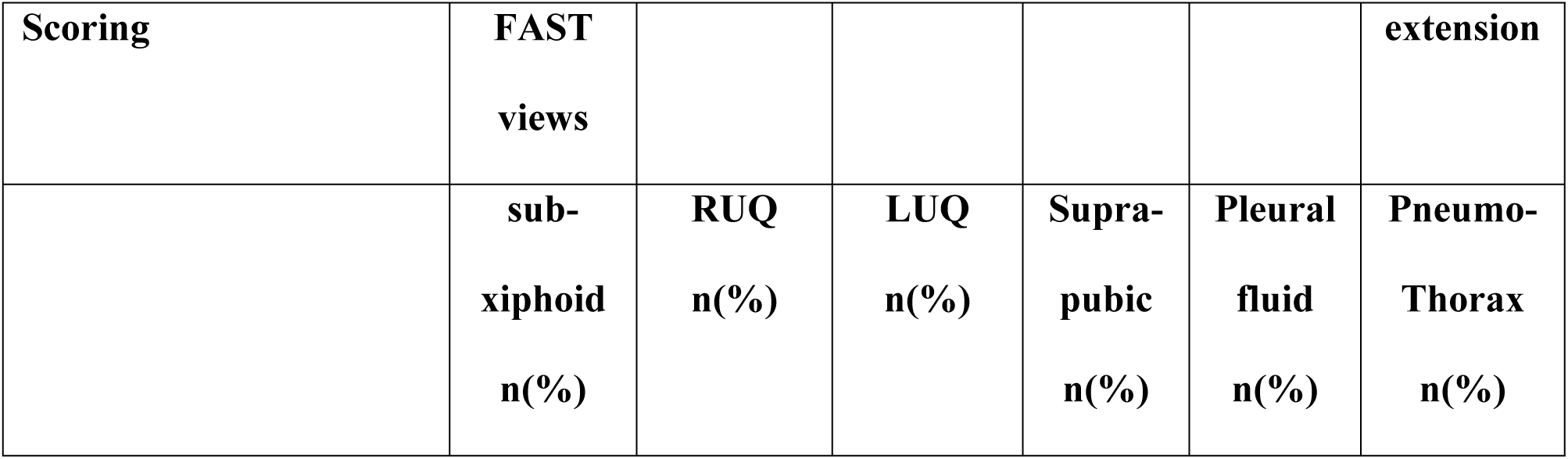
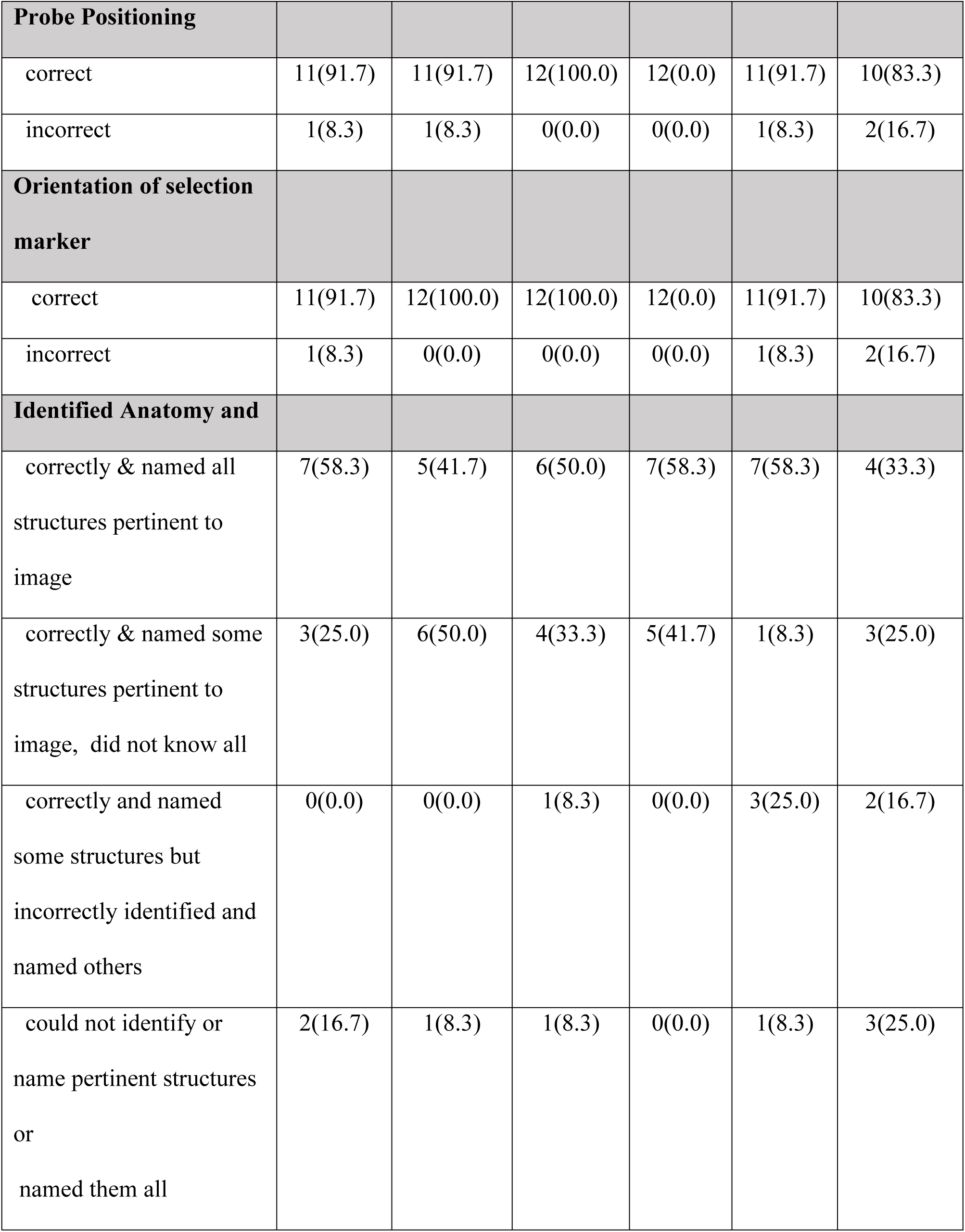
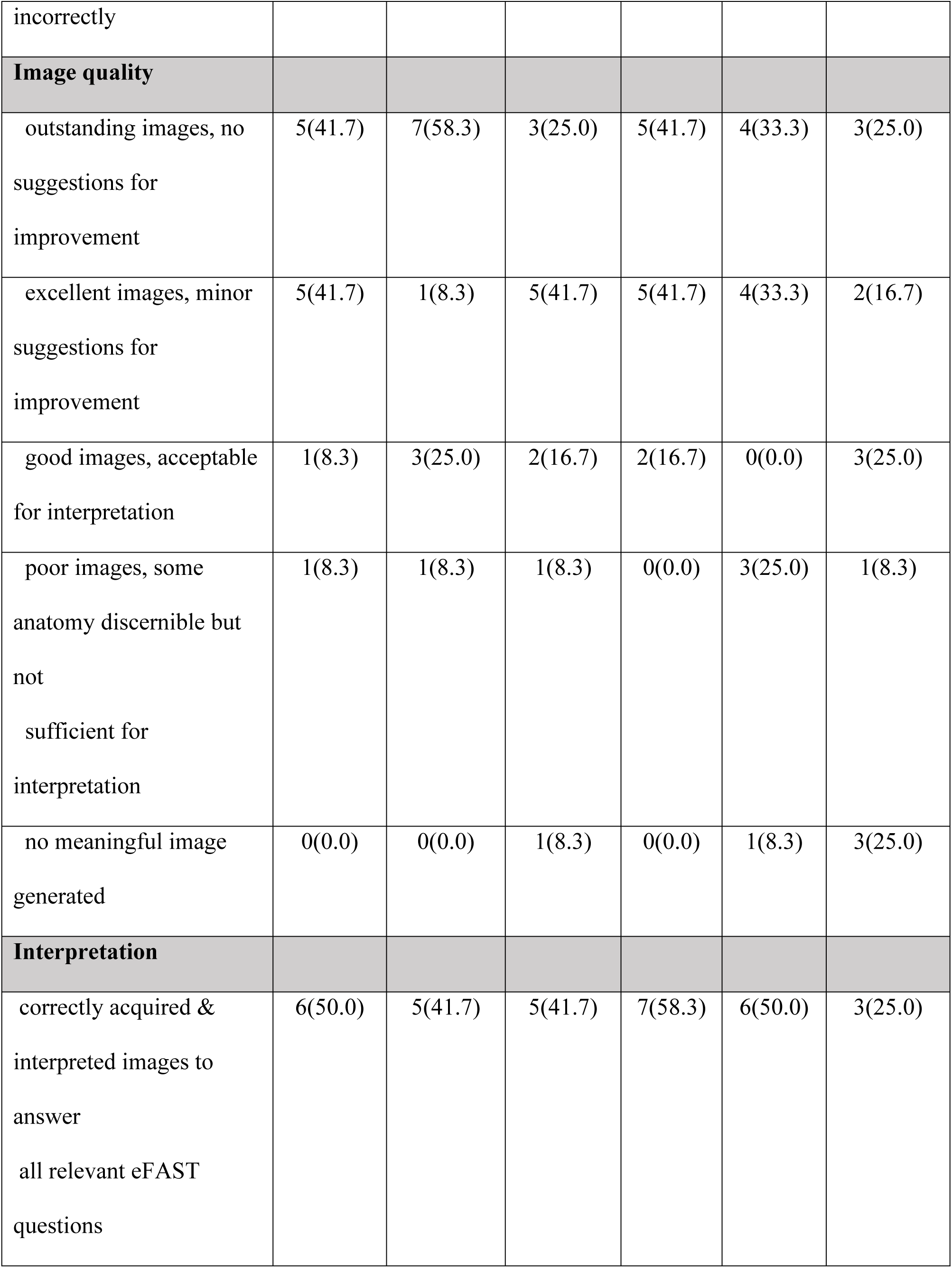
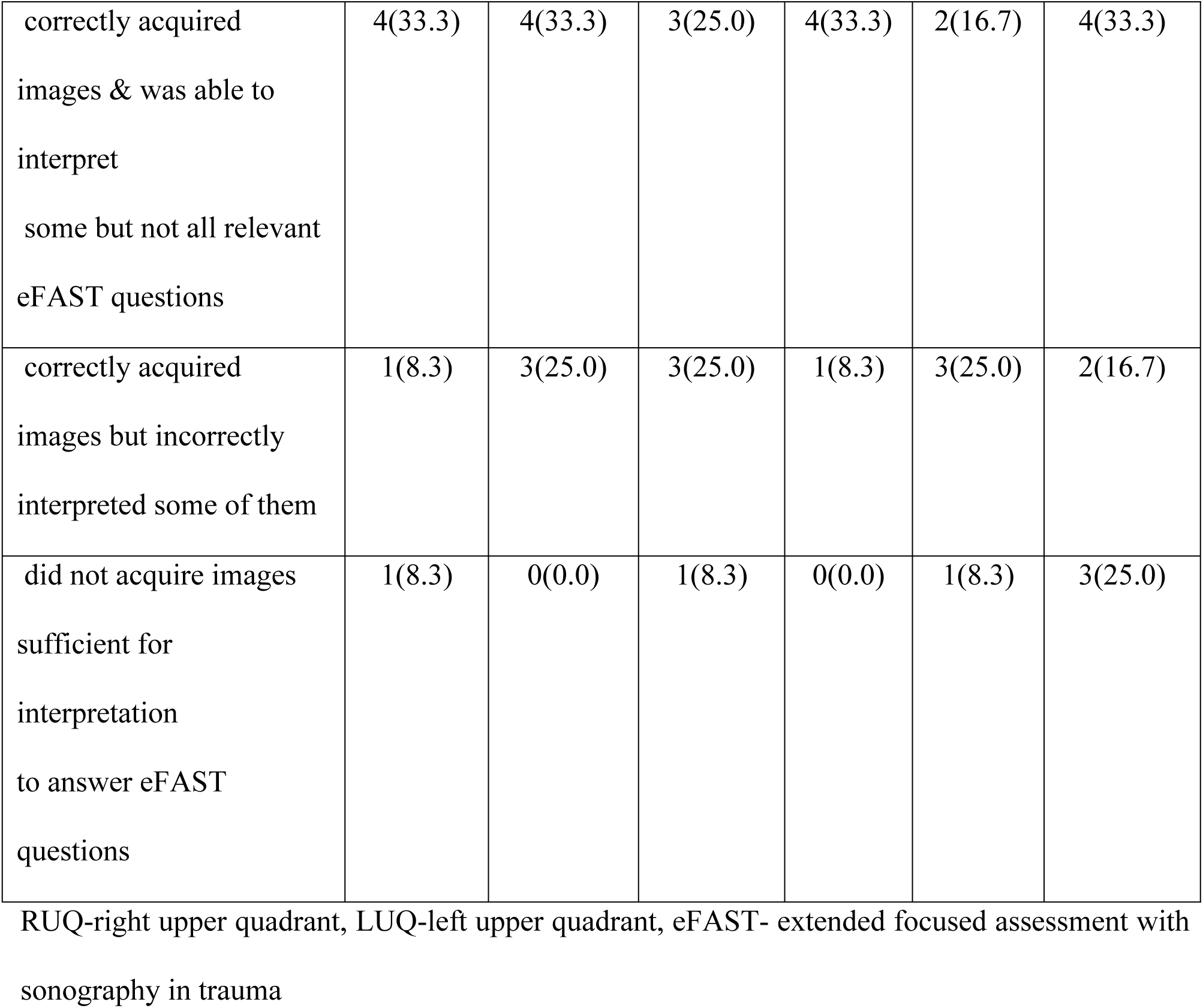
Baseline findings two weeks after training of 12 clinicians on principles of point of care ultrasound in focused assessment with sonography in trauma at Kiwoko hospital, Uganda.

### Follow up after three months

Evaluation of clinicians’ skills in performing eFAST at three months of follow up was good in almost all the eFAST views assessed. However, a non-statistically significant decline (p=0.444) was noted in which 4(44.4%) incorrectly positioned the probe on the right upper quadrant and on the pneumothorax eFAST view.

In addition, at three months follow up, the majority of the clinicians did not obtain outstanding images with no suggestions for improvements in almost all the five eFAST views, but rather obtained excellent images with minor suggestions for improvement in each of the five eFAST views. For instance, in sub-xiphoid view, none of them obtained outstanding quality images with no suggestions for improvement, but 6(66.7%) of them obtained quality excellent images with minor suggestions for improvement in the same eFAST view. This observation (decline from baseline findings) however was not statistically significant (p=1.000). A statistically significant decline from baseline findings (p=0.001) in image quality was noted in evaluation of organs in LUQ eFAST view. It was noted that 3(33.3%) clinicians did not generate meaningful images in LUQ and 4(44.4%) generated images that were good and acceptable for interpretation.

At three months follow up, the clinicians correctly acquired and interpreted images to answer all relevant eFAST questions in all the five eFAST views. No significant decline was noted but rather a slight non statistically significant (p>0.05) improvement. Table 3 summarizes this evaluation at three months follow-up assessment of the trainees.

**Table 3:**
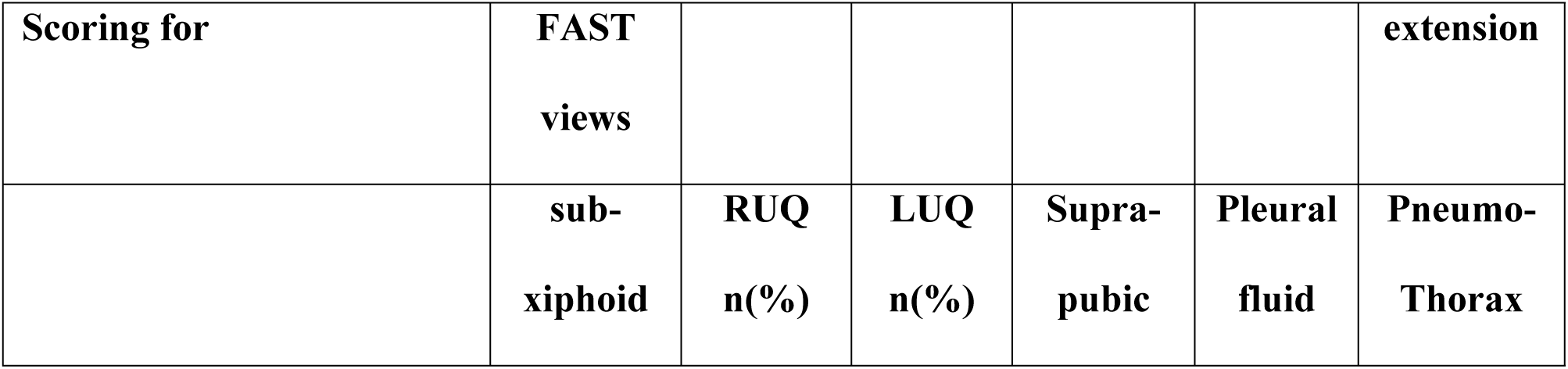
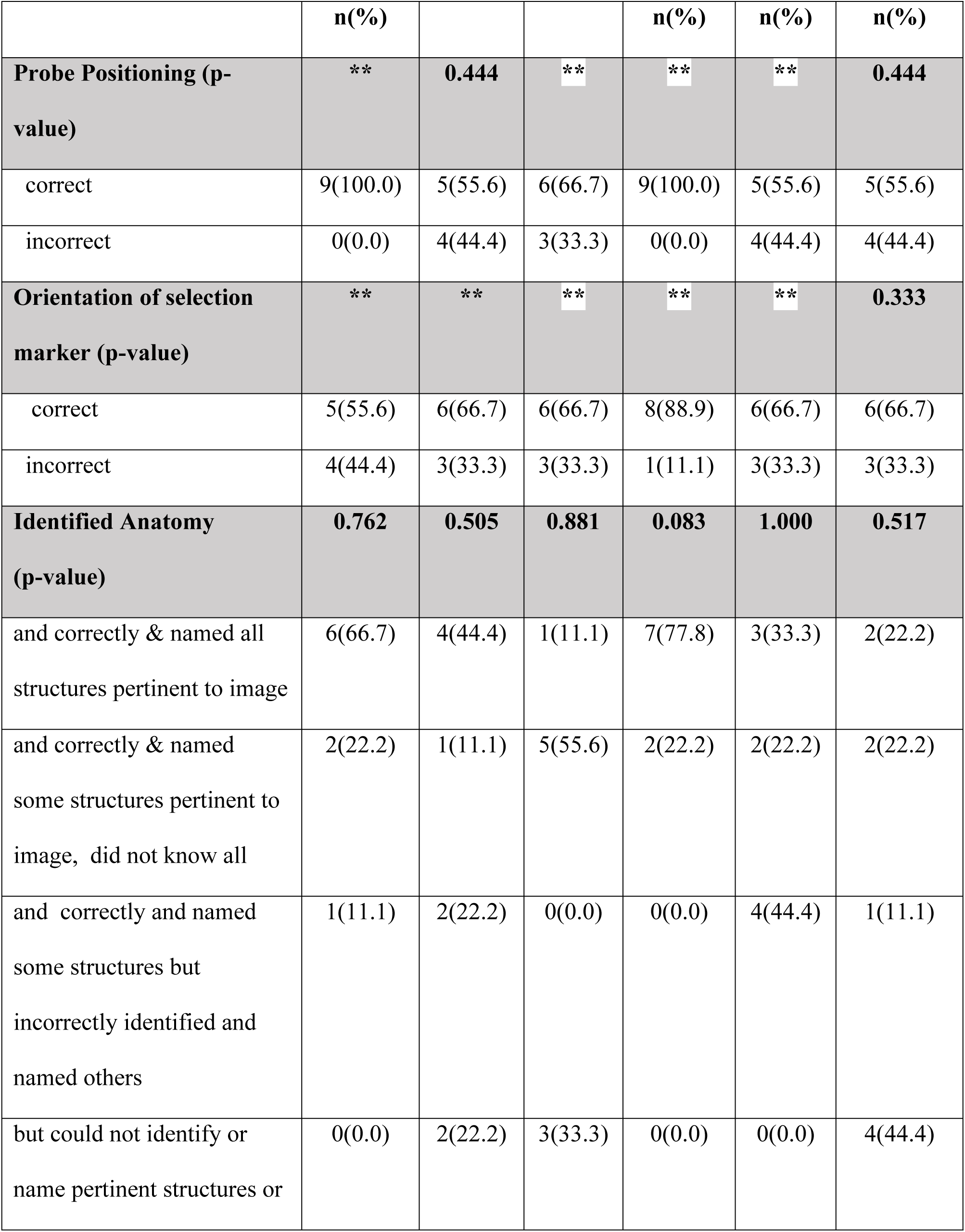
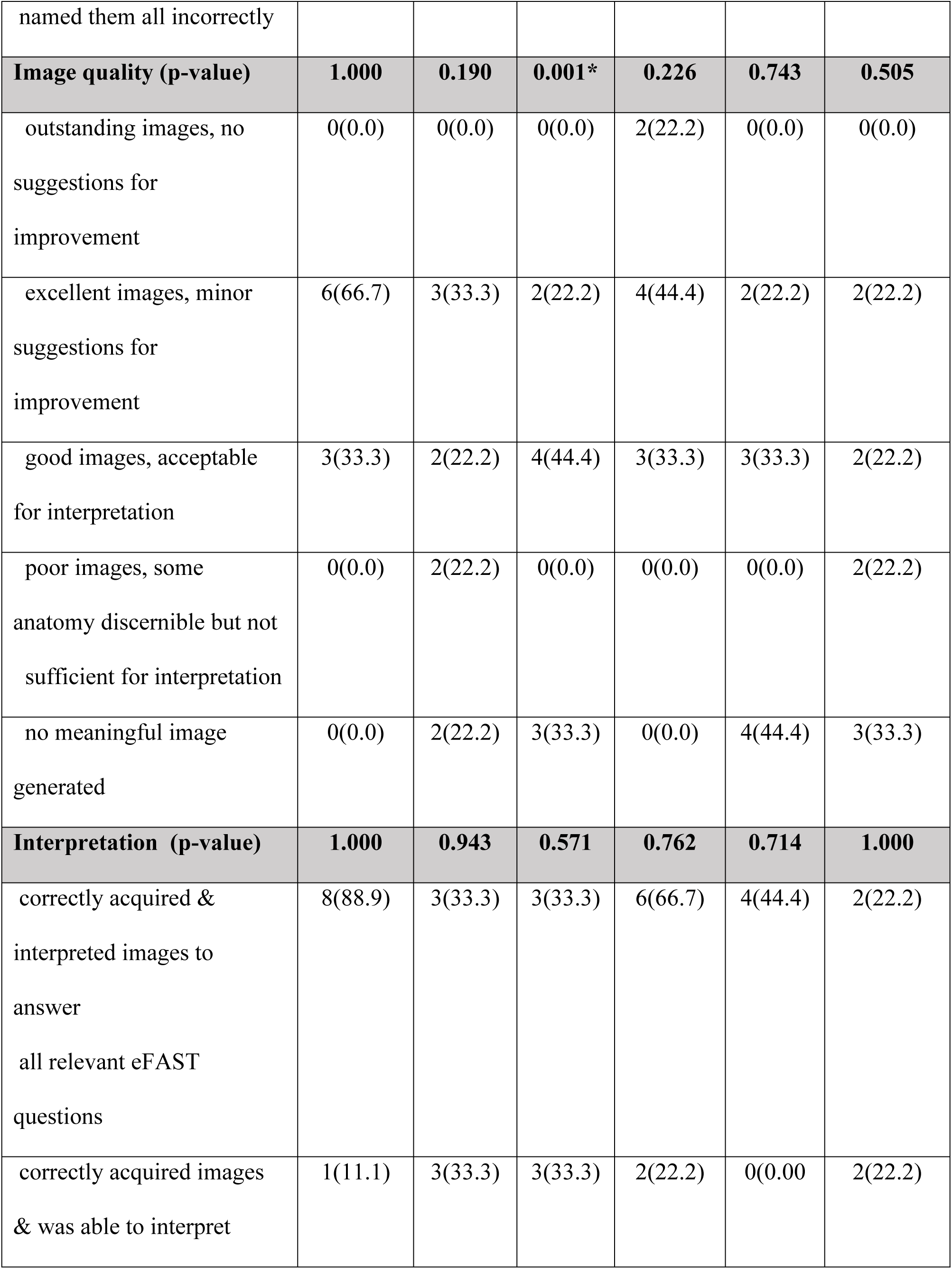
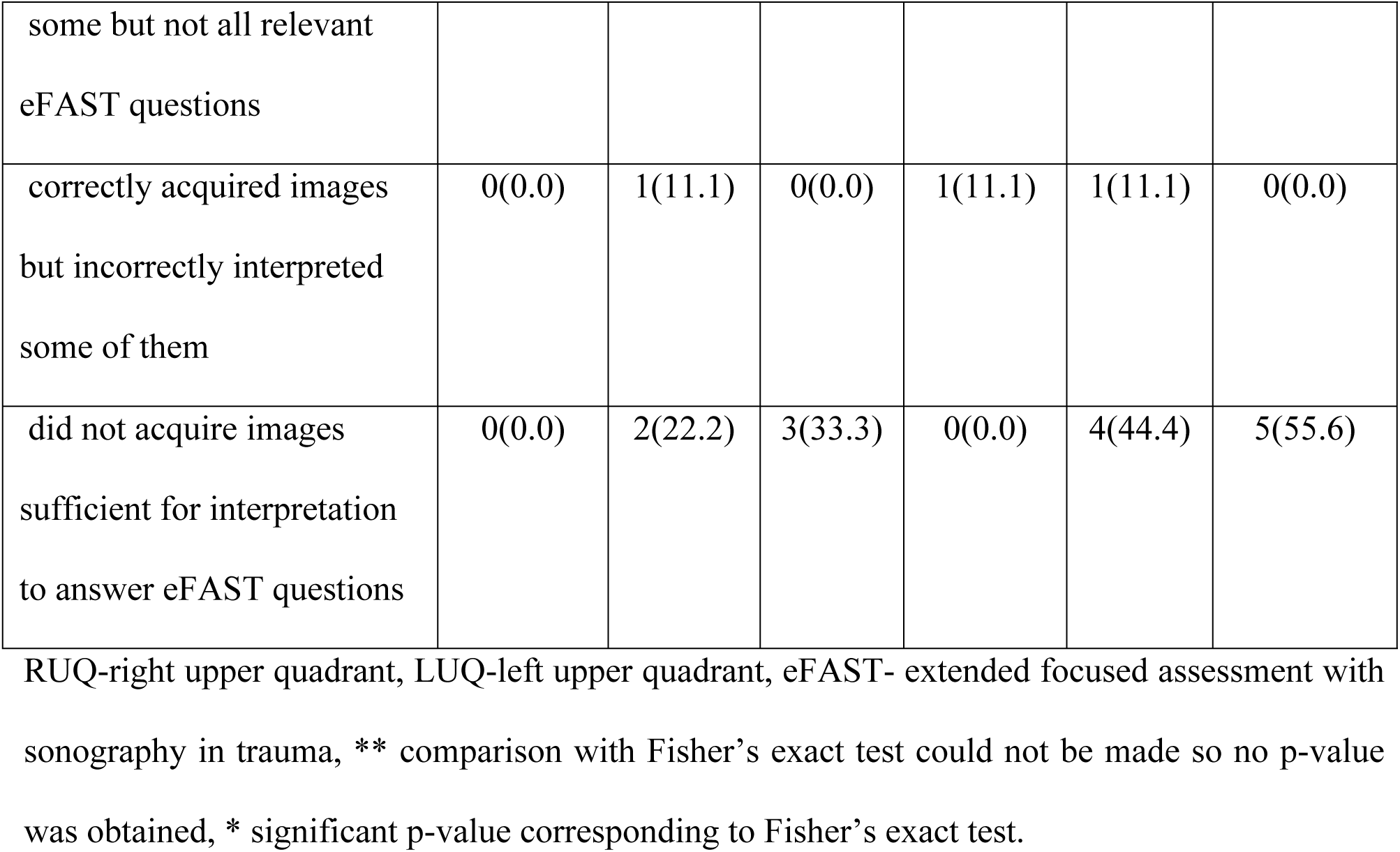
Three months follow-up findings after training of clinicians on principles of point of care ultrasound in focused assessment with sonography in trauma at Kiwoko hospital, Uganda.

### Clinicians’ use of ultrasound scan during three months follow up period

From Table 4, it is evident that 4(44.4%) of the clinicians had performed the ultrasound scan procedure during the follow up period. It also shows that the use of ultrasound changed clinical patient management in 3(33.3%) of the clinicians more than half of the time. From the same table, one can observe that in the three months follow up period, three of the four scans that clinicians performed were in the second month of follow up and the 4^th^ scan was in either the 3^rd^ or the 1^st^ month of follow up.

**Table 4:**
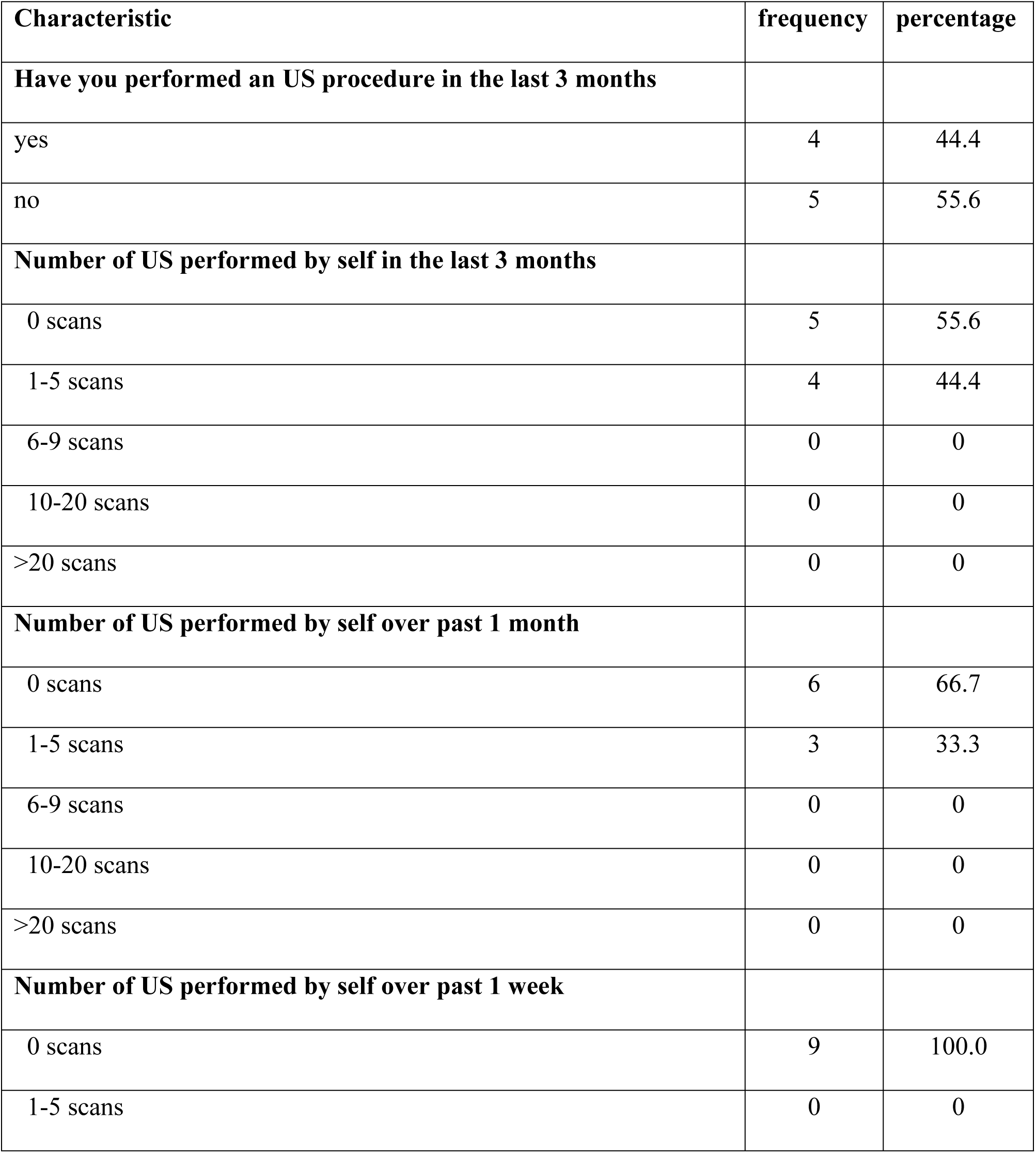
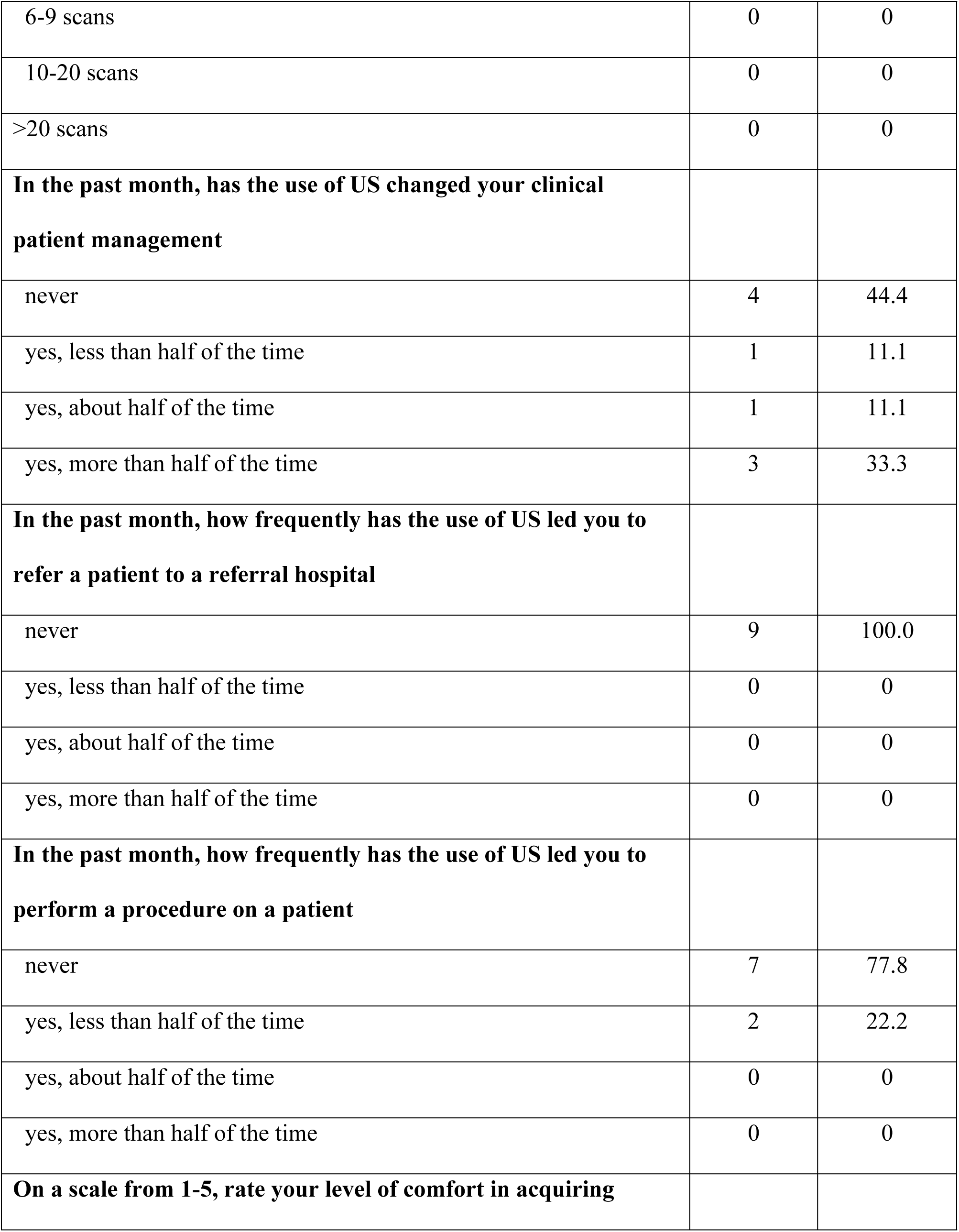
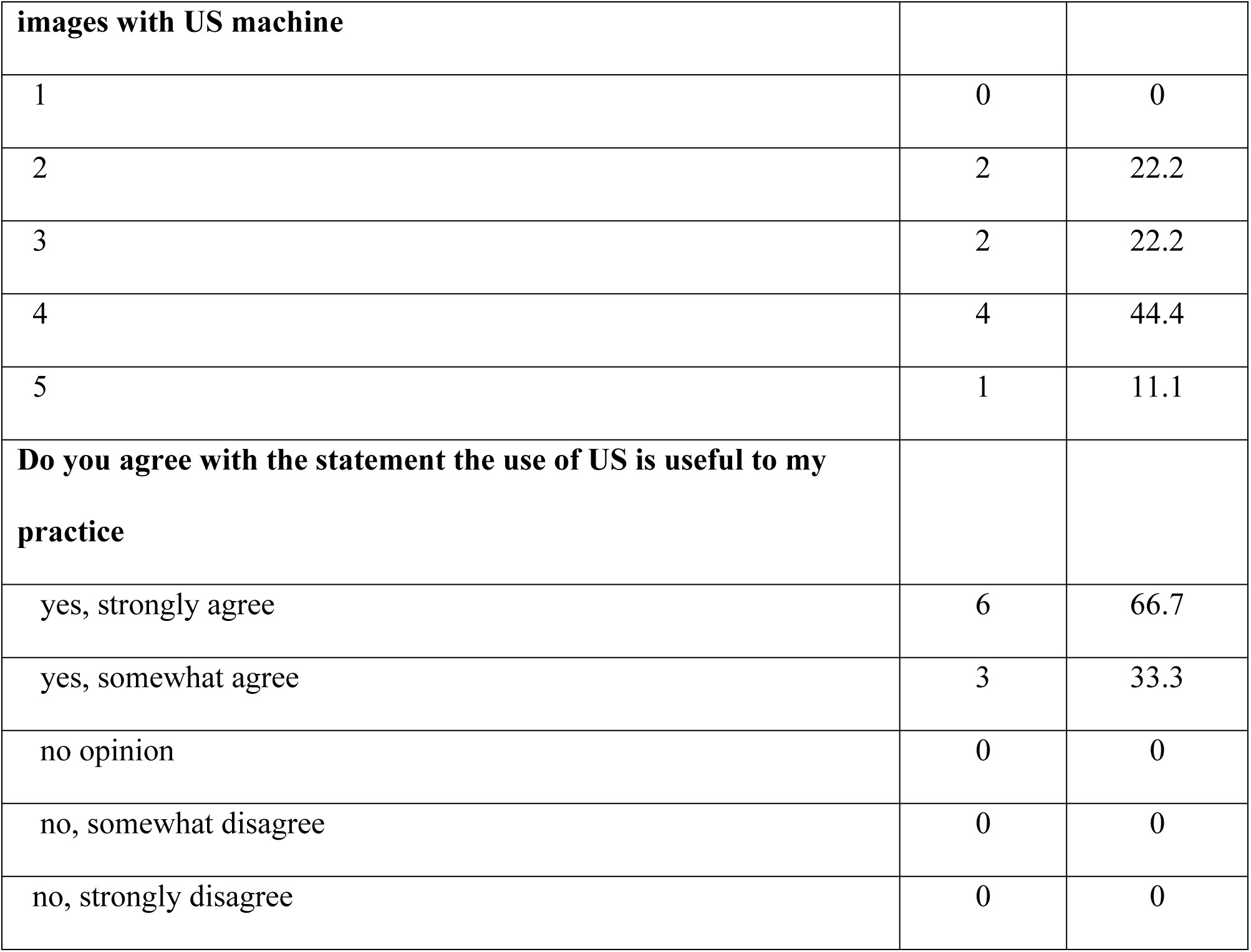
Clinicians practical experience in use of ultrasound scan after training on principles of point of care ultrasound in focused assessment with sonography in trauma at Kiwoko hospital, Uganda.

## Discussion

The purpose of this study was to evaluate the feasibility of implementing point of care ultrasound training in trauma for non-imaging health professionals and evaluate the ability of the trainees to apply the knowledge and skills gained to perform the ultrasound examination in trauma patients. This study brought out strengths and limitations of the eFAST PoCUS training program as well as opportunities for further improvement reported in other literature as well.

The baseline assessment was conducted two weeks post training. At baseline assessment, there was an overall outstanding performance of over 90% in transducer positioning and orientation whereas anatomy, image quality and interpretation of findings were performed averagely with the best performance in suprapubic, followed by sub-xiphoid, pleural fluid, and RUQ views. The LUQ and pneumothorax views had the least performance with scores of as low as 25% in image interpretation and quality of image. This finding is in agreement with a previous study which trained paramedics where the best performance was in the suprapubic, RUQ and pneumothorax views but on the contrary, the least performance was in the subxiphoid view[10, 11]. However another study on eFAST demonstrated poor performance by clinicians in the evaluation of pneumothorax using ultrasound[11]. Our findings may not be surprising given that trainees generally performed well in most areas because they had just been trained and therefore, the knowledge gained was presumably still fresh in their minds. The least performance scored in LUQ and pneumothorax views could possibly be explained by the fact that even in normal imaging practice by qualified imaging experts, these views usually require adequate time and rigorous practice to perform well. Therefore, when it comes to health workers with no imaging background at all, such a performance may not be surprising. Our overall baseline scores was 64% compared to a Kenyan National pilot to train clinicians, where the baseline score was 70% with a 10% improvement in practical and written assessments within a space of five months[12]. Another study to train final year medical students had a baseline score of 88%[13] while a similar study for rural healthcare providers similar to our setting had a baseline score of 74%[13].

Assessment at three months follow up also demonstrated the best performance to be in suprapubic and subxphoid views with scores of 71% and 62% respectively while the least performance was similar to baseline with lowest scores of 36% and 33.4% respectively in the LUQ and pneumothorax views. On the contrary, in a study by Duke et al who trained paramedics, the follow up conducted at two months reported that the best performance was in the RUQ and pneumothorax views and the least performance was in the subxiphoid and suprapubic views[9] We also noted that there was an overall decline of 26.2% in all the parameters assessed with a 22% decline in probe positioning, 27% decline in orientation, 15% decline in anatomy, and 90% decline in image quality but there was an 8.7% improvement in image interpretation whereas PoCUS study in kenya had an improvement of 5% in the test scores at three and six months[14].

Assessment of clinicians after three months showed that approximately 50% of the clinicians actually performed eFAST ultrasound on their own following the training compared to 85% use of eFAST POCUS by interns in the emergency department in Thailand.

In this study, we generally note that trainees had lower scores than what has been reported in other literature. The possible explanation for this is not very clear-cut, however, this could possibly be due to the nature of the training, duration of the training as well as competing work demands of the trainees. All our trainees were still full time employees expected to attend to their duty in the hospital and as such they had limited time to concentrate and there was no dedicated time for training. This forced them to be in and out of training as they had to attend to patients which ultimately affected their concentration. Second, we had shorter training times and the trainees had few contact hours with the trainers when compared to other studies reported. It is thus most likely that the limited contact time with trainers did not avail the trainees with enough practical exposure. This observation however brings out significant pointers as we move into implementing eFAST POCUS among non-imaging health workers. First, for someone to competently perform eFAST to acceptable levels, there must be enough time set aside to train to gain both knowledge and skills. Second, contact supervision with a qualified imaging expert for a prolonged period of time is more likely to result in better eventual competency and proficiency when compared to shorter training periods that are also affected by other competing work demands. Therefore, settings where eFAST POCUS is being implemented need to think about these observations and ensure that there is adequate training time, support supervision and protected time for trainees.

A number of studies have reported about the success of eFAST and POCUS, mainly from high income countries[10, 11, 13]. This study conducted in Uganda contributes to this body of knowledge from the context of low-resource settings. The fact that we implemented this POCUS study at a rural based hospital with high volumes of patient numbers and very few health workers is a strength of the study as it demonstrates that POCUS in trauma patients can be implemented if the necessary resources and time are available. The trainees in this study demonstrated some level of retention of knowledge and skills post training and with support supervision and follow up training, such trainees can attain acceptable levels of competency. Few trainees were retained for subsequent follow up which is a major limitation of the study, but it also highlights the challenges low-resource settings are likely to encounter when implementing POCUS because of high turnover and thus the remaining few get overwhelmed. In this study, we did not intend to conduct a highly powerful bed-side skills assessment with high psychometric rigor involving a number of trauma cases. We thus encourage further research in areas implementing POCUS to conduct rigorous longitudinal skills assessments for both knowledge and skills retention at the bedside.

## Conclusion

This study has demonstrated that with adequate training and support supervision, clinicians without prior exposure to ultrasound can perform the eFAST exam and recognize abnormal fluid collections and pathology in trauma patients. Although we noted a decline at follow-up in image quality and low performance in the LUQ and pneumothorax views, overall, the trainees that were followed up demonstrated acceptable retention of basic knowledge and skills. There is need for continuous training, regulation and support supervision to improve efficiency in areas of poor performance. The training has the potential to improve eFAST knowledge and skills amongst clinicians in rural areas if adequate resources such as longer contact hours, support supervision and protected time to train are in place. The utility of eFAST POCUS among clinicians should be further advocated for and supported.

## Data Availability

All relevant data are within the manuscript and its Supporting Information files.

